# Comparative Performance of Claude and GPT Models in Basic Radiological Imaging Tasks

**DOI:** 10.1101/2024.11.16.24317414

**Authors:** Cindy Nguyen, D Carrion, MK Badawy

**Affiliations:** Department of Medical Imaging and Radiation Sciences, Monash University, Clayton, VIC, 3800, Australia; Monash Imaging, Monash Health, Clayton, VIC, 3168, Australia

## Abstract

**Background:** Publicly available artificial intelligence (AI) Visual Language Models (VLMs) are constantly improving. The advent of vision capabilities on these models could enhance workflows in radiology. Evaluating their performance in radiological image interpretation is vital to their potential integration into practice.

**Aim:** This study aims to evaluate the proficiency and consistency of the publicly available VLMs, Claude and GPT, across multiple iterations in basic image interpretation tasks.

**Method:** Subsets from publicly available datasets, ROCOv2 and MURAv1.1, were used to evaluate 6 VLMs. A system prompt and image were inputted into each model thrice. The outputs were compared to the dataset captions to evaluate each model’s accuracy in recognising the modality, anatomy, and detecting fractures on radiographs. The consistency of the output across iterations was also analysed.

**Results:** Evaluation of the ROCOv2 dataset showed high accuracy in modality recognition, with some models achieving 100%. Anatomical recognition ranged between 61% and 85% accuracy across all models tested. On the MURAv1.1 dataset, Claude-3.5-Sonnet had the highest anatomical recognition with 57% accuracy, while GPT-4o had the best fracture detection with 62% accuracy. Claude-3.5-Sonnet was the most consistent model, with 83% and 92% consistency in anatomy and fracture detection, respectively.

**Conclusion:** Given Claude and GPT’s current accuracy and reliability, integration of these models into clinical settings is not yet feasible. This study highlights the need for ongoing development and establishment of standardised testing techniques to ensure these models achieve reliable performance.

## Introduction

The integration of artificial intelligence (AI) models into the medical domain is already becoming a reality in contemporary Australian healthcare with acknowledgement from professional bodies such as the Australian Health Practitioner Regulation Agency (APHRA)[1] and the Royal Australian and New Zealand College of Radiologists (RANZCR) [2]. Radiology, as a data-driven specialty, can greatly benefit from these models in both interpretive and non-interpretive uses, from report generation to streamlining administrative processes [3], [4]. Task-specific specialised models are already being seen in clinical practice, such as those that can detect radiologic findings on chest X-rays [5] or do real-time detection and triaging of cerebral haemorrhage cases on CT [6].

While these task-specific models are beneficial, they are inflexible and can only perform tasks predefined by their training dataset [7]. As such, there has been a turn towards developing generalist foundation models that can perform a diverse range of tasks with little to no additional training [7]. Advancements in multimodal architectures and self-supervised learning that allow for training on large amounts of unlabelled data have made this possible [7], [8]. Successful development and validation of foundation models will reduce the need for multiple task-specific models, which has the potential to simplify the deployment and management of AI systems [9]. While medical foundation models are in development, in August 2024, Zhang et al. [9] touted their BiomedGPT model as the first fully transparent generalist medical foundation model to be thoroughly evaluated. However, research and attention to foundation models have preceded this, with many looking at publicly available large language models (LLMs) such as OpenAI’s GPT due to its accessibility and ease of use [10], [11].

In 2022, the release of ChatGPT showcased the model’s in-context learning abilities, allowing it to learn from a limited number of examples and generalise to new test sets [7]. The model was exclusively text-based at this stage and lacked image interpretation capabilities. By early February 2023, the first article exploring the application of ChatGPT in radiology was published, followed by over 50 additional articles within six months[12]. These studies predominantly addressed non-image analysis tasks such as generating differential diagnoses from text prompts, structured reporting, and enhancing patient communication [12]. In September 2023, the GPT model range was extended to include vision capabilities with the introduction of GPT-4V, transforming it into a vision-language model (VLM) capable of processing textual and visual data. This advancement prompted rapid evaluation of the model’s image interpretation abilities, potentially expanding its utility in clinical settings. However, initial assessments revealed that the vision capabilities were not yet reliable for clinical use, performing below the standards of its text-based counterpart [13], [14].

Despite the limitations in their image interpretation capabilities, existing studies have highlighted the substantial potential of these models. In March 2024, Anthropic introduced the Claude 3 model family, followed by an update to Claude Sonnet in June 2024, positioning them as direct competitors to OpenAI’s GPT models [15]. These models possess similar multimodal capabilities, prompting researchers to rapidly publish evaluations of Claude’s performance compared to GPT-4 [16], [17]. However, the volume of studies assessing the image interpretation abilities of Claude models remains significantly lower than those focusing on GPT models, likely due to the recent release of Claude.

This study seeks to contribute to the developing body of research by providing one of the initial evaluations of the image interpretation capabilities of Claude models relative to GPT models. Demonstrating satisfactory performance in fundamental tasks could facilitate the integration of these models into radiologic workflows, such as patient triage and screening, and potentially enhance the radiographer’s ‘Red Dot System’—a method in which radiographers annotate radiographs suspected of containing abnormalities [18]. Additionally, accuracy in image interpretation is critical for more advanced applications, including automated report generation. The primary objective of this study is to evaluate the proficiency and consistency of Claude and GPT models in essential diagnostic tasks, explicitly focusing on accurately identifying imaging modalities, anatomical regions, and fractures in radiographic images.

## Method

### Dataset Construction

In this retrospective study, two publicly available datasets were used: ROCOv2 to assess the models’ proficiency in recognising image modalities and anatomical regions [19] and MURAv1.1 to evaluate proficiency in identifying anatomical regions and detecting fractures in upper appendicular radiographs [20].

ROCOv2 is a large manually validated dataset extracted from the PMC Open Access Subset that covers a range of modalities, anatomical regions and medical concepts [19]. A subset of 100 images spans three modalities, including computed tomography (CT), magnetic resonance imaging (MRI) and general X-ray. A breakdown of the anatomical regions included per imaging modality is shown in Table 1. The ROCOv2 dataset also has the limitation of lower image quality. All cases were manually screened and excluded if they had poor image quality or additional annotations on the image to reduce the chances of confounding the results. The ground truth for modality and anatomical region were extracted from the CSV metadata attached to the dataset.

**Table 1.** Aggregated data of anatomical regions by imaging modalities for ROCOv2.

| Modality and anatomical region | | Total number of images $n$ |
| --- | --- | --- |
| Total |  | 100 |
| CT |  | 40 |
|  | Abdomen and pelvis | 15 |
|  | Chest | 14 |
|  | Brain | 10 |
|  | Spine | 1 |
| MRI |  | 31 |
|  | Brain | 15 |
|  | Spine | 16 |
| X-ray |  | 29 |
|  | Chest | 10 |
|  | Abdomen | 3 |
|  | Skeletal | 16 |

MURAv1.1 is a large dataset containing upper extremity musculoskeletal radiographs manually labelled as normal or abnormal. From this, a subset of 300 images were selected across six anatomical regions: elbow, finger, forearm, hand, humerus, and wrist. Images were chosen to ensure a fracture prevalence of 50%, resulting in 25 normal and 25 abnormal images for each region. MURAv1.1 only labelled images as normal or abnormal, encompassing fractures and other abnormalities such as hardware, lesions, degenerative joint diseases, etc. As the scope of this study only included fracture detection, images with a low level of complexity, defined as those with clear and direct radiologic signs, were manually selected to avoid inaccuracies in the ground truth as best as possible.

#### Testing

This study utilised six model variations of Claude and GPT: Anthropic’s Claude 3 Haiku, Claude 3 Opus, and Claude 3.5 Sonnet, and OpenAI’s GPT-4 Turbo, GPT-4o Mini, and GPT-4o. Each of these models underwent independent testing using Python with OpenAI’s (GPT) and Anthropic’s (Claude) APIs to automate the submission of images with the prompt. A system prompt was crafted to guarantee consistent replies for the automated assessment. The system prompt for MURAv1.1 and ROCOv2 can be found in Table 2.

**Table 2.** System prompts for MURAv1.1 and ROCOv2.

| MURAv1.1 Prompt | ROCov2 System Prompt |
| --- | --- |
| You are a useful research assistant. | You are a useful research assistant. |
| You are being provided with images to determine the anatomical region and fracture present. | You are being provided with images to determine the anatomical region and modality. |
| You should return a one-line answer in CSV format and never return header. | You should return a one-line answer in CSV format and never return header. |
| You should NEVER return "Unknown" and instead provide best guess. | You should NEVER return "Unknown" and instead provide best guess. |
| You should return 1st col as Anatomical Region and 2nd col as Fracture present where 1 is YES and 0 is NO. | You should return 1st col as Modality and 2nd col as Anatomical Region. |
| Example output format:<br>Foot,0<br>Back,0<br>Shoulder,1 | Example output format:<br>CT,Brain<br>X-ray,Foot<br>MRI,Brain |

Chat interfaces are nondeterministic, with intentional randomness to mimic natural language. This can result in different outputs generated for the same prompt [21]. Using the API, the temperature hyperparameter can be adjusted to limit randomness; however, to see the standard performance of these models, we opted to use the default values for GPT and Claude, which were 0.7 and 1, respectively. Given this, each test was conducted three times to evaluate consistency and uniformity across several iterations.

To evaluate the models’ performance, the output was compared to the captions collected with the images to determine performance in recognition of imaging modality, anatomical region and detection of fractures. As the focus was on an objective evaluation method, no follow-up questions or avenues for the models to provide reasoning was provided reasoning, such as free text or bounding boxes, were permitted.

### Data Analysis

The primary metric used to evaluate the performance was the accuracy of each model in recognising imaging modality, anatomical region, and detecting fractures in the first iteration. For the ROCOv2 subset, metrics were calculated per modality, while the MURAv1.1 subset was calculated per anatomical region with additional sensitivity and specificity calculations for fracture using Excel. Consistency was defined as having the same answer across all three iterations. Using GraphPad Prism 10, the Chi-squared test was performed to compare models and evaluate the statistical significance using a *p*-value of less than 0.05.

## Results

### Models’ performance in imaging modality and anatomical region identification

The ROCOv2 subset consisted of 100 images distributed between 3 imaging modalities (CT, MRI, X-ray) and various anatomical regions. Several models obtained a 100% (100/100) accuracy rate for modality identification with outstanding consistency (Figure 1). The exceptions were Claude 3 Haiku and Claude 3 Opus, which misidentified CT images as MRI.

**Figure 1.**
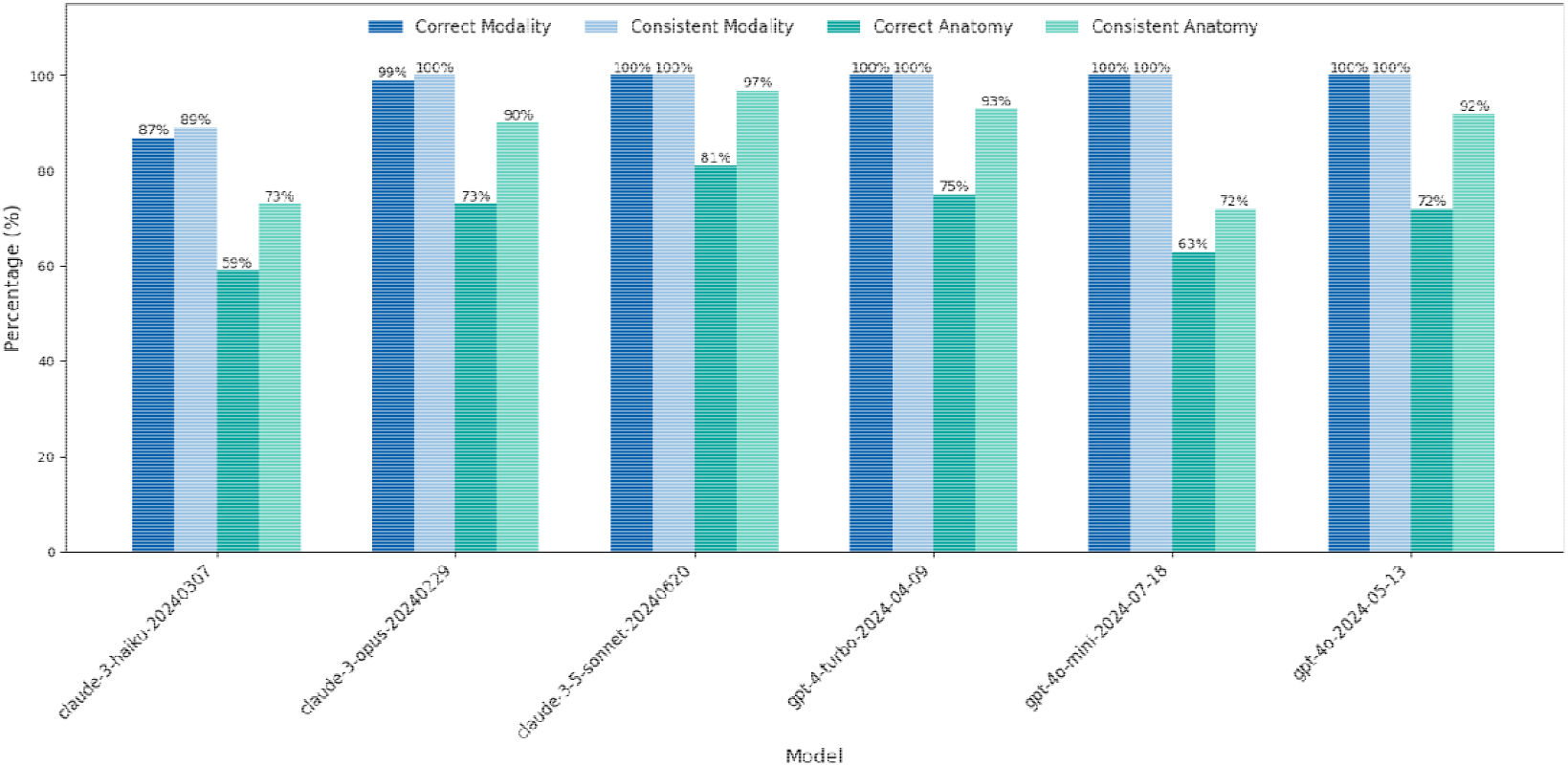
Accuracy and consistency of GPT and Claude models in detecting modality and anatomy from ROCOv2 dataset.

There were varying degrees of success in anatomical region identification. Claude 3.5 Sonnet demonstrated the highest accuracy overall with 85% (85/100) correctly identified, followed by GPT-4 Turbo, which had 78% (78/100) accuracy. Claude 3 Haiku performed the worst overall, with a 61% (61/100) accuracy rate. Generally, the anatomical regions in CT were best recognised, followed by X-ray and then MRI. Claude 3 Opus, Claude 3 Sonnet, GPT-4 Turbo and GPT-4o demonstrated consistent answers in over 90% of cases (Figure 1).

### Models’ performance in anatomical region identification and fracture detection of upper appendicular radiographs

The MURAv1.1 subset consisted of 300 images, with 50 images across six anatomical regions and a 50% fracture prevalence. The anatomical areas included were the elbow, finger, forearm, hand, humerus, and wrist. Overall accuracy and consistency in anatomical region identification and fracture detection are displayed in Figure 2. Across all models, there was poor anatomical identification, with the highest accuracy rate being 57% (171/300) from Claude 3.5 Sonnet. A breakdown of accuracy in anatomical region per region is shown in Table 3.

**Table 3.**
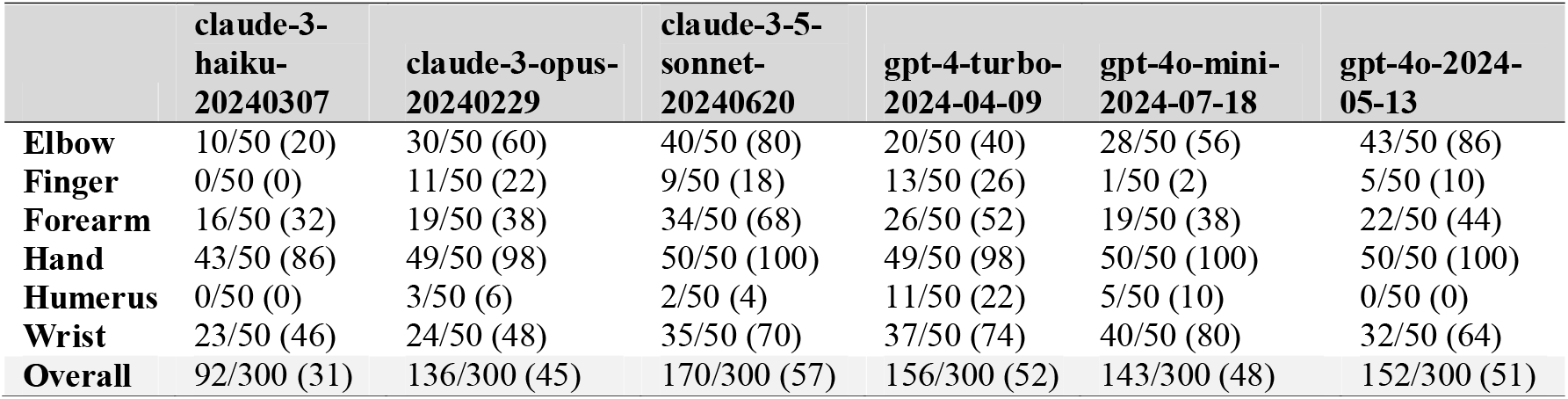
Anatomical region accuracy per region on MURAv1.1 – identified/total (%)

**Figure 2.**
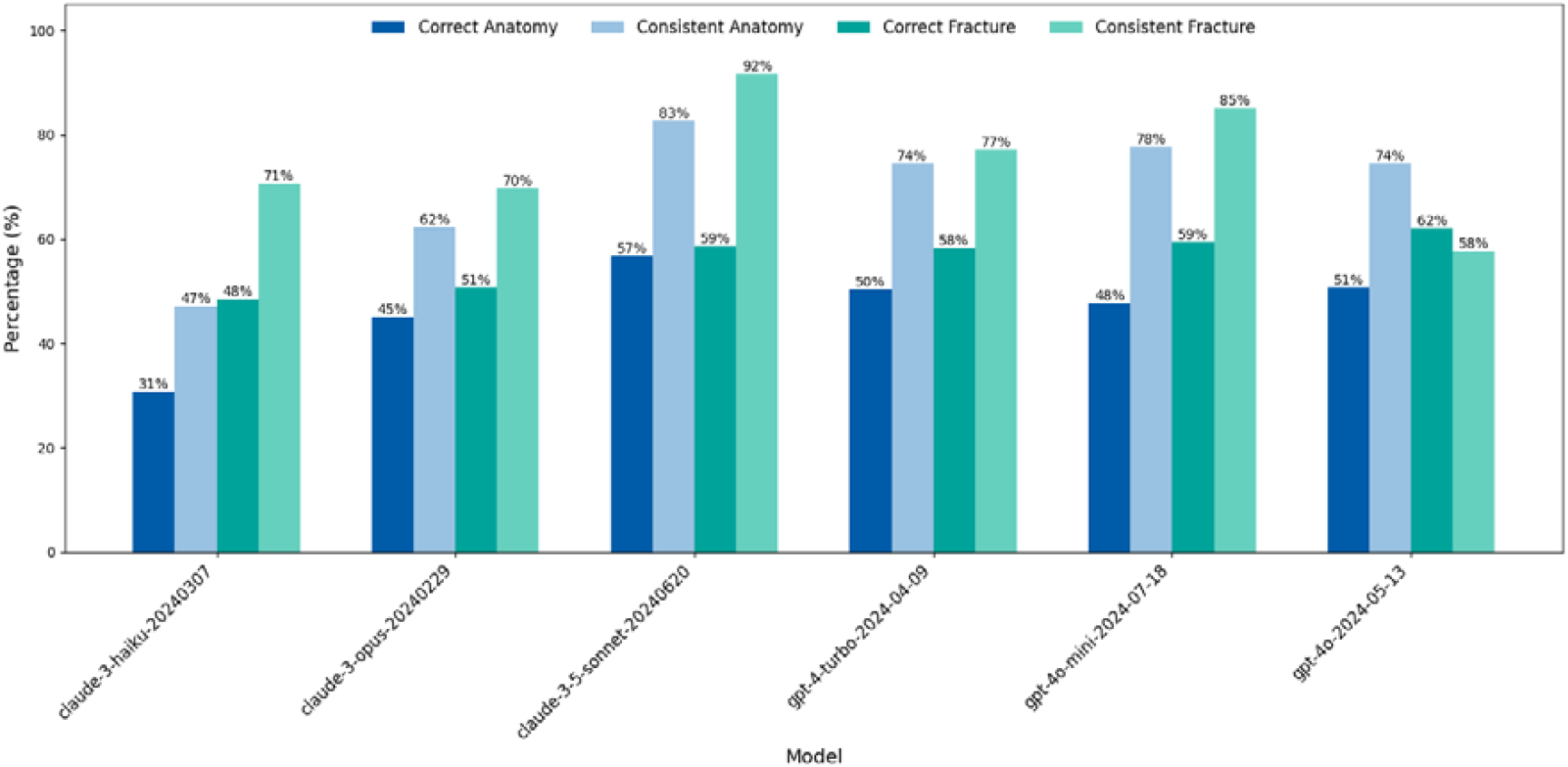
Accuracy and consistency of Claude and GPT models in identifying anatomical regions and fractures from the MURAv1.1 dataset.

Overall, GPT-4o performed the best in detecting fractures with a 62% (186/300) accuracy rate. The next best models were Claude 3.5 Sonnet (difference: 3%, χ2 = 11.9, p = 0.0005) and GPT-4o Mini (difference: 3%, χ2 = 46.8, p = 0.0001). Claude 3.5 Sonnet, however, marked 85% (254/300) of cases as normal, resulting in 114/150 false negatives with a sensitivity of 24% and specificity of 93%. The radar graphs (Figure 3) highlight GPT-4o’s balance across accuracy, sensitivity, and specificity, particularly excelling in detecting fractures across all regions, while the confusion matrices (Figure 4) reveal a high number of false negatives for Claude 3.5 Sonnet, especially in the hand and wrist regions, which directly impacted its sensitivity.

**Figure 3.**
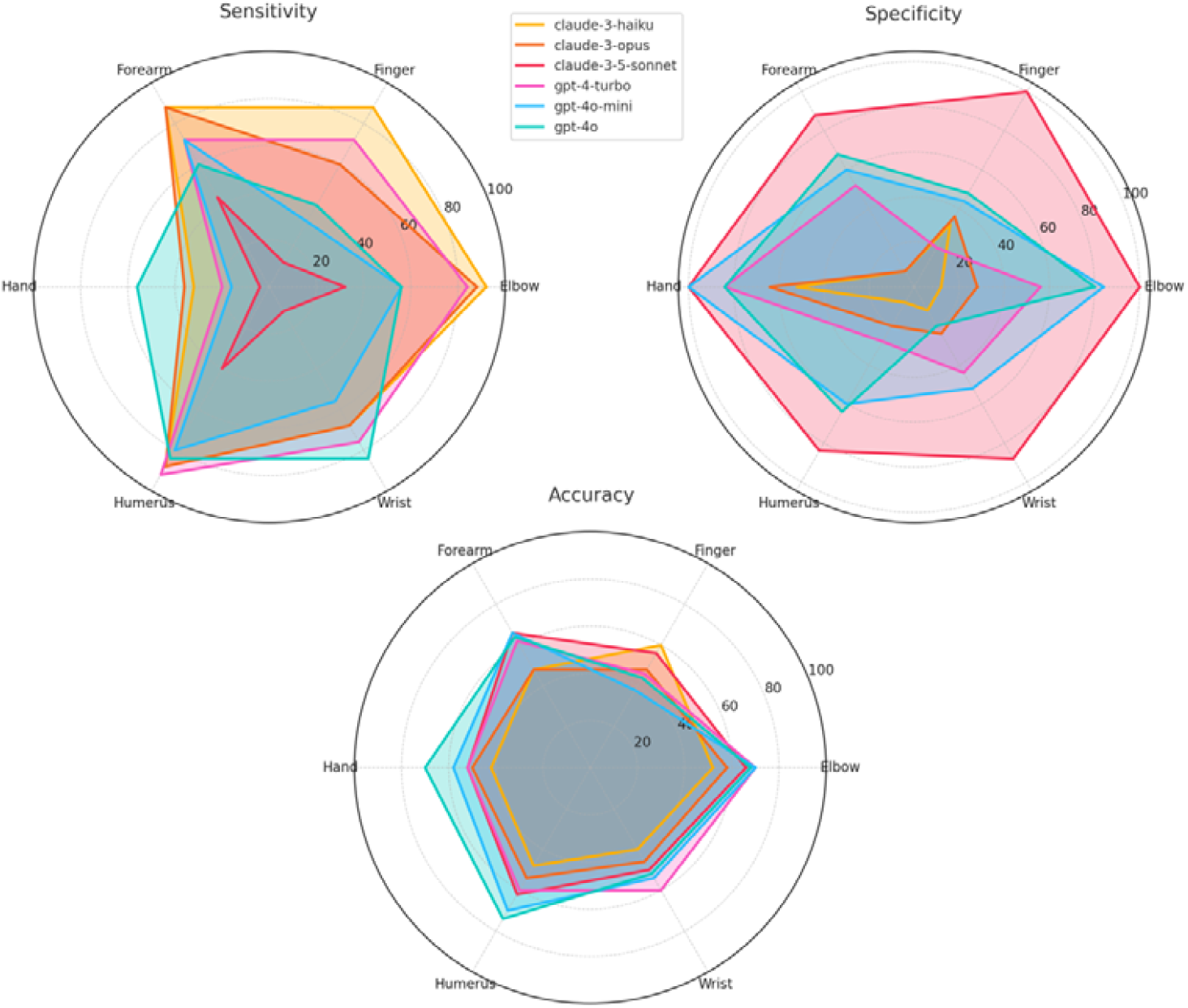
Radar charts illustrating the accuracy, sensitivity, and specificity of each model across different anatomical regions.

**Figure 4.**
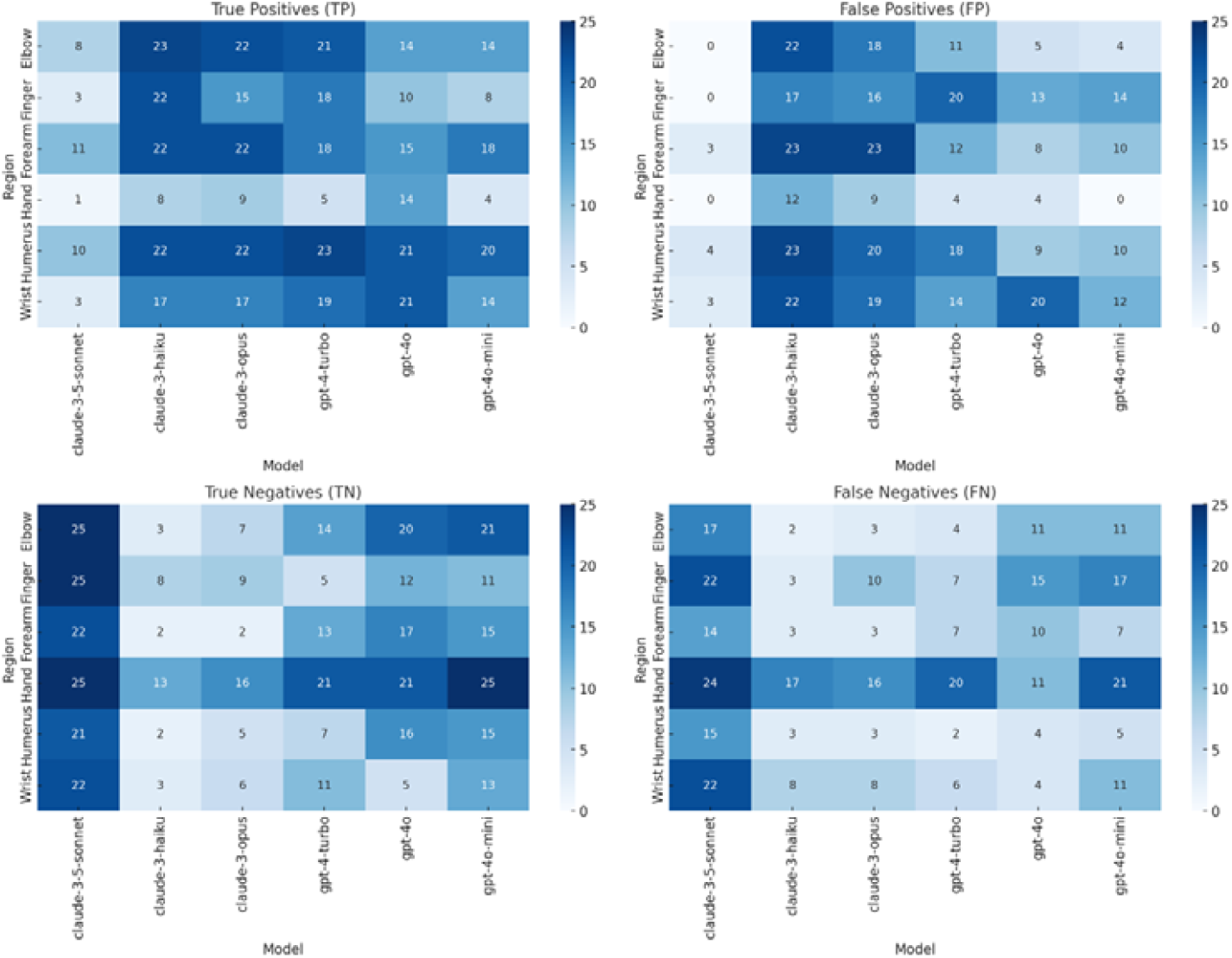
Heatmaps of confusion matrix components (True Positives, False Positives, True Negatives, and False Negatives) across models and regions.

## Discussion

This study provided an objective preliminary evaluation of the Claude-3 and GPT-4 variant models using the identification of imaging modality, anatomical region and detection of fractures to depict the models’ performance in basic radiologic image interpretation tasks. While these models demonstrated that they could identify features on medical images, overall performance in clinically relevant tasks indicates that they cannot be relied on for stand-alone radiological interpretation.

On the ROCOv2 data subset, there was a satisfactory performance in the accuracy and consistency of the models, with many achieving 100% accuracy for imaging modality identification and at least 70% for the anatomical region. Models could identify the anatomical region on CT images well; however, they fell short for MRI images, with accuracy ranging from only 42% –74%. This outcome was likely because the models did not specify the level for lumbar spine images and predicted only ‘Spine’ in all but one case across all models. Given that lumbar spine images accounted for 26% (8/31) of all MRI cases, this was detrimental to their accuracy outcome on anatomical region identification. The answer of ‘Spine’ was not accepted because the same error was not encountered for cervical spine images to the same degree. Although both tasks are considered rudimentary with little clinical value, most models performed well, demonstrating that they can identify relevant features in medical images.

Model performance in the MURAv1.1 data subset focussing on upper appendicular radiographs was substantially lower. Claude 3.5 Sonnet’s identification of anatomical region on the MURAv1.1 data set had an accuracy of 57% (170/300). While this was the highest-performing model on the MURAv1.1 dataset, this accuracy rate was still lower than the lowest-performing model on the ROCOv2 dataset – Claude 3 Haiku with 61% accuracy (61/100). These two subsets are not directly comparable as there is a difference in the number and the modality of the cases. However, when we isolate the x-ray images from the ROCOv2 subset, the lowest-performing model – Claude 3 Haiku with 65.5% accuracy (19/29) – still outperformed the highest-performing model on the MURAv1.1 dataset. The main regions of difficulty were finger and humerus studies. Finger studies were often predicted as ‘Hand’ while humerus studies were often predicted as either ‘Elbow’, ‘Shoulder’, or ‘Arm’. This may be because the predicted anatomical regions are components of the actual anatomical region, and the models did not consider the whole image.

Fracture detection on the MURAv1.1 subset is where there is more clinical relevance. Fracture detection is one of the most common radiologic tasks in trauma patients, often with clear radiologic signs. Despite this, the models did not perform well in this task, with the best accuracy outcome from GPT-4o with 62% (186/300) correctly identified cases. However, Claude 3.5 Sonnet was quite consistent with 83% and 92% consistency for the anatomical region and fracture detection across the three iterations, indicating reliable model performance.

The results of this study are consistent with published literature. Brin et al. [14] evaluated GPT-4 on pathological cases across multiple modalities, finding an overall diagnostic accuracy of only 35.2% with a high rate of hallucinations. Similarly, Reith et al. [17] evaluated GPT-4, Gemini-1.5-Pro and Claude 3 Opus on paediatric radiological images and found an overall accuracy of 27.8%. Kurokawa et al. [16], comparing Claude 3 Opus and Claude 3.5 Sonnet, and Horiuchi et al. [13], comparing GPT-4-based ChatGPT and GPT-4V-based ChatGPT, both found that there was poorer accuracy on image-based questions than on text-only questions. However, models designed specifically for fracture detection perform quite well, with Guermazi et al. [22] finding that the stand-alone performance of their model had an AUC of 0.97. This highlights the current difference in the efficacy of specialised models and publicly available VLMs. Despite this, the ability to integrate both textual and visual data more closely aligns with the decision-making process of radiologists, meaning that VLMs have the potential to surpass task-specific image analysis models mainly based on convolution neural networks [14].

As one of the aims of this study was to provide an objective evaluation, we did not explore avenues that would give insight into the model’s output. Studies that did use free-text outputs could comment on more qualitative findings revealing that hallucinations, where the models provide incorrect or fabricated information, were quite common, constituting a large limitation to future integration [14], [17]. In addition, prompting models to produce an ‘explanation’ for its output does not reflect the mechanical process underlying the prediction. It can, therefore, be used only to promote critical thinking rather than understanding the model itself [23]. Without insight into the models’ structure, training data or model weights, we cannot explain the behaviour and outputs of an AI model [23]. Explainable AI is, therefore, another field of interest in AI development as it can increase physician and patient trust in these models.

Quality benchmark datasets can also increase the trustworthiness of AI models [24]. Reliable benchmark datasets can be used as standardised tests, which enable more consistent and objective evaluation and comparison of AI systems. However, benchmark creation is not an easy process with many considerations, such as the representatives of cases, quality of labelling, image quality and de-identification [24]. Our method in this study could contribute to a more comprehensive and unified framework for benchmarking and standardised testing. Strides have already been made with RadBench from Wu et al. [25].

Our study had some limitations. As these models are trained on a proprietary mix that includes publicly available information on the Internet [15], we cannot be sure that the ROCOv2 and MURAv1.1 datasets were not included in the training set, which could lead to an overestimation of model performance. There is potential selection bias due to the subjective selection of cases. Furthermore, images with no fractures were selected from the cases in MURAv1.1 that were labelled as negative for abnormalities; however, this was not verified. Image interpretation performance was not evaluated with the textual clinical context provided. Doing so could have impacted model performance positively as it would more closely align with real decision-making processes. Our study had a limited scope, mainly focussing on fractures in upper appendicular radiographs; however, this is not fully representative of the range of pathology investigated with medical imaging. Finally, these models are constantly updating, so the results found in this study only represent a snapshot of time.

## Conclusion

In this study, the current performance in accuracy and reliability of Claude and GPT demonstrates that integration of these models into clinical settings is not yet feasible. This study highlights the need for ongoing development and establishment of standardised testing techniques to ensure these models achieve reliable performance. Further works should evaluate the performance of Claude and GPT on a wider range of pathologies and images that are not publicly available.

## Data Availability

All data produced in the present study are available upon reasonable request to the authors.

## References

[1] Australian Health Practitioner Regulation Agency, “Meeting your professional obligations when using Artificial Intelligence in healthcare.” Accessed: Nov. 09, 2024. [Online]. Available: https://www.ahpra.gov.au/Resources/Artificial-Intelligence-in-healthcare.aspx

[2] The Royal Australian and New Zealand College of Radiologists, “Generative Artificial Intelligence and Large Language Models: Position Paper.” Accessed: Nov. 08, 2024. [Online]. Available: https://www.ranzcr.com/our-work/artificial-intelligence

[3] T. Nakaura et al., “The impact of large language models on radiology: a guide for radiologists on the latest innovations in AI,” Jpn J Radiol, vol. 42, no. 7, pp. 685–696, Jul. 2024, doi: 10.1007/s11604-024-01552-0.

[4] C. Mello-Thoms and C. A. B. Mello, “Clinical applications of artificial intelligence in radiology,” The British Journal of Radiology, vol. 96, no. 1150, p. 20221031, Oct. 2023, doi: 10.1259/bjr.20221031.

[5] Y. Vasilev, A. Vladzymyrskyy, O. Omelyanskaya, I. Blokhin, Y. Kirpichev, and K. Arzamasov, “AI-Based CXR First Reading: Current Limitations to Ensure Practical Value,” Diagnostics, vol. 13, no. 8, p. 1430, Apr. 2023, doi: 10.3390/diagnostics13081430.

[6] L. Li et al., “Deep Learning for Hemorrhagic Lesion Detection and Segmentation on Brain CT Images,” IEEE J. Biomed. Health Inform., vol. 25, no. 5, pp. 1646–1659, May 2021, doi: 10.1109/JBHI.2020.3028243.

[7] M. Moor et al., “Foundation models for generalist medical artificial intelligence,” Nature, vol. 616, no. 7956, pp. 259–265, Apr. 2023, doi: 10.1038/s41586-023-05881-4.

[8] R. Krishnan, P. Rajpurkar, and E. J. Topol, “Self-supervised learning in medicine and healthcare,” Nat. Biomed. Eng, vol. 6, no. 12, pp. 1346–1352, Aug. 2022, doi: 10.1038/s41551-022-00914-1.

[9] K. Zhang et al., “A generalist vision–language foundation model for diverse biomedical tasks,” Nat Med, vol. 30, no. 11, pp. 3129–3141, Nov. 2024, doi: 10.1038/s41591-024-03185-2.

[10] H. Kim, P. Kim, I. Joo, J. H. Kim, C. M. Park, and S. H. Yoon, “ChatGPT Vision for Radiological Interpretation: An Investigation Using Medical School Radiology Examinations,” Korean J Radiol, vol. 25, no. 4, p. 403, 2024, doi: 10.3348/kjr.2024.0017.

[11] A. Lecler, L. Duron, and P. Soyer, “Revolutionizing radiology with GPT-based models: Current applications, future possibilities and limitations of ChatGPT,” Diagnostic and Interventional Imaging, vol. 104, no. 6, pp. 269–274, Jun. 2023, doi: 10.1016/j.diii.2023.02.003.

[12] K. Bera, G. O’Connor, S. Jiang, S. H. Tirumani, and N. Ramaiya, “Analysis of ChatGPT publications in radiology: Literature so far,” Current Problems in Diagnostic Radiology, vol. 53, no. 2, pp. 215–225, Mar. 2024, doi: 10.1067/j.cpradiol.2023.10.013.

[13] D. Horiuchi et al., “ChatGPT’s diagnostic performance based on textual vs. visual information compared to radiologists’ diagnostic performance in musculoskeletal radiology,” Eur Radiol, Jul. 2024, doi: 10.1007/s00330-024-10902-5.

[14] D. Brin et al., “Assessing GPT-4 multimodal performance in radiological image analysis,” Eur Radiol, Aug. 2024, doi: 10.1007/s00330-024-11035-5.

[15] Ahtropic, Claude. [Online]. Available: https://www.anthropic.com/claude

[16] R. Kurokawa et al., “Diagnostic performances of Claude 3 Opus and Claude 3.5 Sonnet from patient history and key images in Radiology’s ‘Diagnosis Please’ cases,” Jpn J Radiol, Aug. 2024, doi: 10.1007/s11604-024-01634-z.

[17] T. P. Reith, D. M. D’Alessandro, and M. P. D’Alessandro, “Capability of multimodal large language models to interpret pediatric radiological images,” Pediatr Radiol, vol. 54, no. 10, pp. 1729–1737, Aug. 2024, doi: 10.1007/s00247-024-06025-0.

[18] M. B. O’Leary et al., “Diagnostic accuracy of radiographers’ red dot usage in the emergency department,” Journal of Radiation Research and Applied Sciences, vol. 17, no. 1, p. 100795, Mar. 2024, doi: 10.1016/j.jrras.2023.100795.

[19] J. Rückert et al., “ROCOv2: Radiology Objects in COntext Version 2, an Updated Multimodal Image Dataset,” Sci Data, vol. 11, no. 1, p. 688, Jun. 2024, doi: 10.1038/s41597-024-03496-6.

[20] P. Rajpurkar et al., “MURA: Large Dataset for Abnormality Detection in Musculoskeletal Radiographs,” May 22, 2018, arXiv: arXiv:1712.06957. Accessed: Nov. 16, 2024. [Online]. Available: http://arxiv.org/abs/1712.06957

[21] J. H. Lee and J. Shin, “How to Optimize Prompting for Large Language Models in Clinical Research,” Korean J Radiol, vol. 25, no. 10, p. 869, 2024, doi: 10.3348/kjr.2024.0695.

[22] A. Guermazi et al., “Improving Radiographic Fracture Recognition Performance and Efficiency Using Artificial Intelligence,” Radiology, vol. 302, no. 3, pp. 627–636, Mar. 2022, doi: 10.1148/radiol.210937.

[23] A. Sarkar, “Large Language Models Cannot Explain Themselves,” May 07, 2024, arXiv: arXiv:2405.04382. Accessed: Nov. 16, 2024. [Online]. Available: http://arxiv.org/abs/2405.04382

[24] N. Sourlos et al., “Recommendations for the creation of benchmark datasets for reproducible artificial intelligence in radiology,” Insights Imaging, vol. 15, no. 1, p. 248, Oct. 2024, doi: 10.1186/s13244-024-01833-2.

[25] C. Wu, X. Zhang, Y. Zhang, Y. Wang, and W. Xie, “Towards Generalist Foundation Model for Radiology by Leveraging Web-scale 2D&3D Medical Data,” 2023, arXiv. doi: 10.48550/ARXIV.2308.02463.

